# Functional Antibodies in COVID-19 Convalescent Plasma

**DOI:** 10.1101/2021.03.08.21253157

**Authors:** Jonathan D. Herman, Chuangqi Wang, Carolin Loos, Hyunah Yoon, Johanna Rivera, M. Eugenia Dieterle, Denise Haslwanter, Rohit K. Jangra, Robert H. Bortz, Katharine J. Bar, Boris Julg, Kartik Chandran, Douglas Lauffenburger, Liise-anne Pirofski, Galit Alter

## Abstract

In the absence of an effective vaccine or monoclonal therapeutic, transfer of convalescent plasma (CCP) was proposed early in the SARS-CoV-2 pandemic as an easily accessible therapy. However, despite the global excitement around this historically valuable therapeutic approach, results from CCP trials have been mixed and highly debated. Unlike other therapeutic interventions, CCP represents a heterogeneous drug. Each CCP unit is unique and collected from an individual recovered COVID-19 patient, making the interpretation of therapeutic benefit more complicated. While the prevailing view in the field would suggest that it is administration of neutralizing antibodies via CCP that centrally provides therapeutic benefit to newly infected COVID-19 patients, many hospitalized COVID-19 patients already possess neutralizing antibodies. Importantly, the therapeutic benefit of antibodies can extend far beyond their simple ability to bind and block infection, especially related to their ability to interact with the innate immune system. In our work we deeply profiled the SARS-CoV-2-specific Fc-response in CCP donors, along with the recipients prior to and after CCP transfer, revealing striking SARS-CoV-2 specific Fc-heterogeneity across CCP units and their recipients. However, CCP units possessed more functional antibodies than acute COVID-19 patients, that shaped the evolution of COVID-19 patient humoral profiles via distinct immunomodulatory effects that varied by pre-existing SARS-CoV-2 Spike (S)-specific IgG titers in the patients. Our analysis identified surprising influence of both S and Nucleocapsid (N) specific antibody functions not only in direct antiviral activity but also in anti-inflammatory effects. These findings offer insights for more comprehensive interpretation of correlates of immunity in ongoing large scale CCP trials and for the design of next generation therapeutic design.

## Introduction

The emergence of SARS-CoV-2 has caused an international pandemic unseen since the 1918 flu^1^. The COVID-19 pandemic has led to worldwide lockdowns, declining economies, overburdened health care centers, and killed over 2 million people ^2,3^. Severe COVID-19, which develops in14% of the infected population^4^, can lead to acute respiratory distress syndrome, renal failure, thromboembolic complications, a hyperinflammatory syndrome, and death^5-7^. While successful vaccine development has progressed at an unprecedented speed ^8^, vaccination continues to roll out slowly, and new viral variants continue to emerge that evade humoral immunity^9-11^, pointing to a continued need for highly effective therapeutics.

Among the therapeutic strategies that gained attention early in the COVID-19 pandemic, COVID-19 convalescent plasma (CCP) was proposed as a possible therapeutic^12^ due to the previous deployment of convalescent plasma in the treatment of the 1918 flu^13^, SARS^14,15^, and H1N1^16,17^. Conceptually, CCP would deliver a bolus of SARS-CoV-2-specific antibodies that would neutralize the virus and blunt viral replication and disease. Now, a year into the COVID-19 pandemic, CCP has been shown to be a safe intervention ^18^, but the therapeutic benefit of this treatment remains unclear.

Randomized control trials suggest that early administration of CCP in COVID-19 is essential for its efficacy^19^. When given within 3 days of symptom onset, high titer CCP resulted in a 73% reduction in the risk of COVID-19 progression^19^. However, when given later in illness, in severely ill COVID-19 patients, no benefit in reducing mortality was observed^20^. Although differences across treated patients may explain some of the inconsistent results observed across trials, the variability of CCP represents one of the key differences in therapeutic impact. Specifically, unlike traditional therapeutics, CCP is not generated from pooled plasma. Rather, each unit of CCP represents a unique collection of antibodies, administered to one or a few COVID-19 patients^21^. Part of the variability of CCP is due to the design of clinical trials. While SARS-CoV-2 titers^19,20,22,23^ or neutralization levels^24^ are used in some trials to guide the selection of CCPs, levels vary across trials and in some instances CCP is used without confirmation of any antibody levels^25,26^. Moreover, each unit of CCP, derived from an individual patient rather than pooled, represents a heterogenous therapy that may complicate interpretation of efficacy of CCP as a therapeutic. Specifically, each CCP unit may differ based on host-genetics^27-29^, severity of antecedent COVID-19 illness^30-32^, and time from recovery from COVID-19 ^32,33^. However, whether CCP heterogeneity extends beyond neutralization and IgG titer and whether particular characteristics of CCP therapy impact the evolution of disease in COVID-19 patients remains unclear but could guide the future interpretation of large scale CCP trials.

Beyond binding and neutralization titers, alterations in pathogen-specific antibody subclass, isotype, Fc-receptor binding, and Fc-effector functions can contribute both directly to pathogen clearance but also indirectly to dampening inflammatory responses^34^. Thus, to begin to define the influence of CCP heterogeneity on recipient humoral immunity, we deeply profiled the SARS-CoV-2 response in units of CCP, as well as in recipients prior to and after CCP transfer. Systems Serology was performed on 19 hospitalized patients with severe COVID-19 disease and the CCP units they received prior to administration. Striking differences were noted in SARS-CoV-2 specific antibody profiles across CCP donors as well as between CCP donors and recipients. Despite the heterogeneity, Fc effector functions were fascinatingly enriched in CCP donors. The level of pre-existing anti-SARS-CoV-2 antibodies determined how CCP-delivered S-specific antibodies either reduced or increased the evolution of inflammatory humoral immune responses in COVID-19 patients. These data point to unexpected effects of CCP on recipient humoral immune responses, pointing to a critical importance of CCP-antibody functional profiles as a potential key metric for the selection and design of highly effective CCP or next generation monoclonal therapeutics.

## Results

### Antibody Profiles of COVID-19 Convalescent Plasma

Mixed results have been reported for clinical trials using CCP to trat COVID-19. When given to patients within 3 days of symptom onset, CCP has stopped progression to severe disease^19^. However, CCP has had more variable effects in the treatment of moderate and severe COVID-19 disease^20,25,26^. Resolution of these often-conflicting results has been difficult due to substantial differences in the criteria used to select CCPs across trials. However, the heterogeneity of each unit of CCP complicates our understanding as well. CCP is a polyclonal mix of antibodies collected from individual donors. If each unit of CCP varies with respect to SARS-CoV-2 specific isotypes, subclass, Fc-receptor binding, and ability to recruit Fc-effector function, that all may collectively contribute to the efficacy of CCP. Given our emerging appreciation for the role of Fc effector functions in both resolution of COVID-19^30^ and vaccine induced immunity^35,36^, here we hypothesized that functional antibody profiles could help resolve the question of CCP’s effect, positive and negative, on COVID-19 disease.

A total of 19 severely ill COVID-19 patients, who received CCP within 72 hours of admission in April and early May 2020, and the CCP they received were profiled in this study **(Supplemental Table 1)**. All patients required supplemental oxygen, had a median WHO ordinal scale value of 5, and 48% of these patients required non-invasive positive pressure ventilation or mechanical ventilation at enrollment. Patients received 1 unit of 200mL CCP pre-screened for Spike IgG titer by ELISA (as described in Yoon et al.^37^) within 3 days of hospitalization (median day 1 of hospitalization) and were followed for up to 28 days after enrollment. Antibody profiling was performed on units of CCP, patients pre-CCP (day −1), one day after CCP (day 1), and 3 days after CCP (day 3). CCP from 14 different donors who recovered from mild to moderate COVID-19 was used in this study. Units from 4 CCP donors were each given to 2 unique patients. CCP units from the 10 other donors were each given to a single patient. The CCP unit could not be identified for one patient. The majority of patients, 94%, were on corticosteroids. Only one patient each received the following other COVID-19 investigational therapies at the time of the trial: remdesivir, sarilumab, and leronlimab. Of the 19 patients, 21% died, 21% remained hospitalized, and 58% were discharged **(Supplemental Table 1)**.

Initial analysis of SARS-CoV-2 specific antibody profiles in CCP units revealed striking yet expected heterogeneity across donor plasma (**Figure 1A**). While Spike (S)-specific IgG1 was present in all units of CCP, substantial variation was noted in IgG-subclass distribution across the units. A subset of CCPs harbored an IgG3-centric response (CCP unit 10 and 14), and a separate subset exhibited an expanded IgA response (CCP unit 6, 7, and 9). Neutralizing antibody titers varied among the donors, with CCP units 12, 13, and 14 having the highest titers. The Fc-receptor (FcR) binding also varied substantially across CCPs, with FCR2A binding expanded in CCP units 7, 10, and 14, with expanded binding to all FcRs in CCP unit 14. The antibody effector profiles evolved proportionally with the overall size of the subclass/isotype/FcR binding profiles, with notable Fc-effector functions present in all units of CCP, albeit the broadest levels of functions were noted in in CCP unit 1, 7, 10, and 14. Only low-level correlations were noted between neutralizing antibody titer and the S-specific Fc profiles across the CCP units (**Figure 1B, Supplemental Figure 1**). Collectively, these data point to striking heterogeneity in CCP profiles that may influence the therapeutic benefit of this polyclonal mix of antibodies.

**Figure 1.**
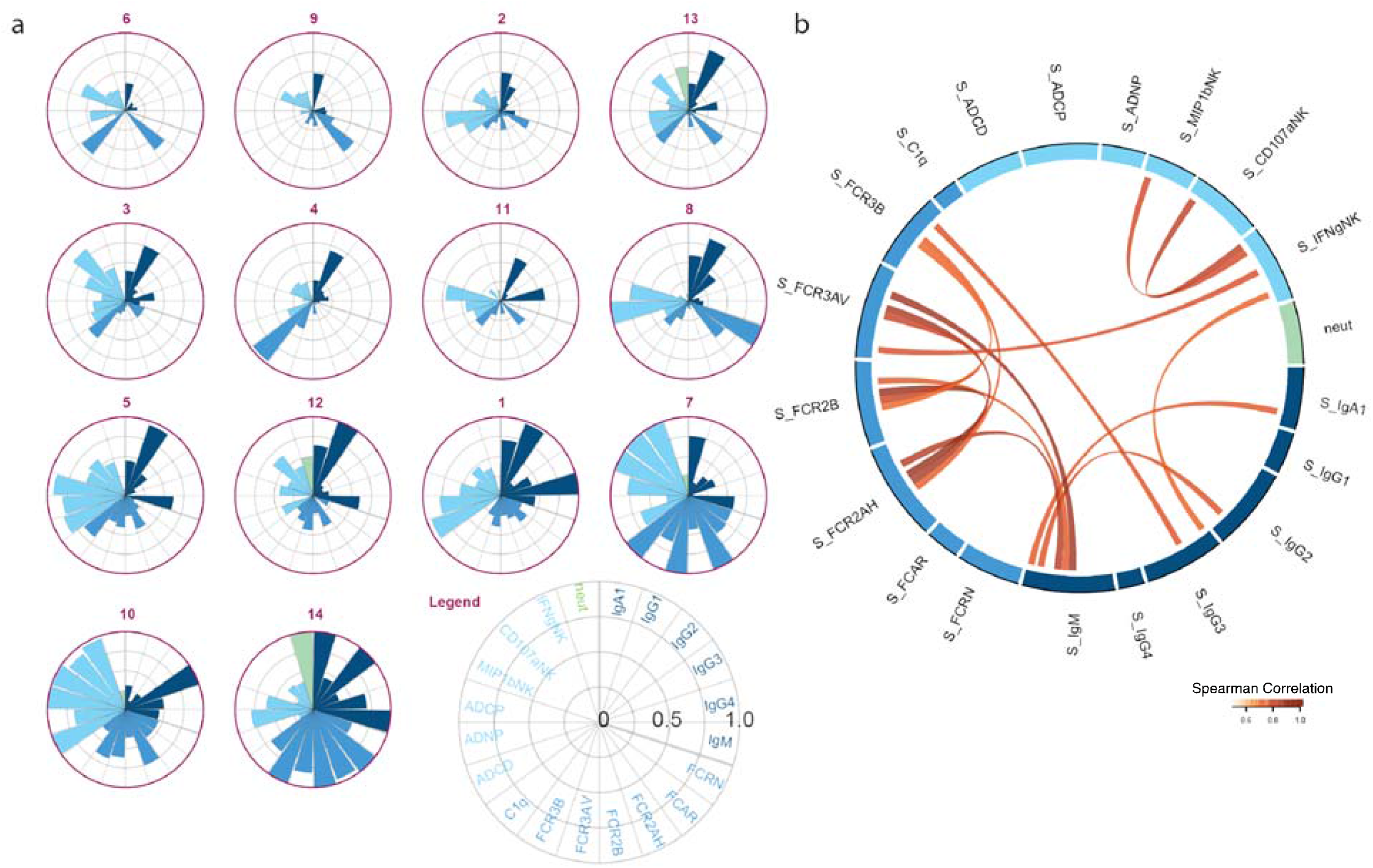
Heterogeneity of Anti-Spike Antibody Profiles in Convalescent Plasma is not Captured by Neutralizing Antibody Titer. Donor CCP were profiled for SARS-CoV-2 Spike-specific antibody responses. (A) Each polar plot depicts an individual donor’s anti-Spike antibody profile, scaled to the minimum and maximum of the 14 units of CCP profiled. Each wedge represents a SARS-CoV-2 antibody feature, and the size of the wedge indicates the magnitude of the value. The colors represent the feature group: dark blue - antibody isotypes and subclasses; blue – Fc-receptor binding levels; light blue – antibody-dependent functions; light green - neutralizing antibody titer. Polar plots were organized by hierarchical clustering of S-specific antibody features. (B) Cord diagram representing the *statistically significant* correlation among Spike-specific antibody features in donor CCP with spearman correlation > *0. 5*. The width of antibody each feature represents the accumulated values of Spearman correlation coefficients of that feature with all other features included in the diagram.

### Donor Convalescent Plasma is Enriched in Anti-SARS-CoV-2 Functional Antibodies Compared to Recipients’ Plasma

As mentioned above, CCP trials have used distinct criteria for the selection of convalescent donors^38^. However, how these CCP units differ from COVID-19 patient recipient antibodies also remains unclear. Thus, we compared the overall antibody profiles in the severe COVID-19 patients pre-CCP (day - 1) to the overall profiles in the CCP units (**Figure 2A**). While heterogeneity was also observed across severely ill COVID-19 recipients (**Supplemental Figure 2A**), an overall expansion of anti-SARS-CoV-2 titers and FcR binding was observed in pre-CCP severe patients compared to units of CCP, the latter drawn from convalescent, less-inflamed individuals (**Figure 2A**). Specifically, IgA titers and Fcα-receptor (FCAR) binding were most significantly expanded in severe COVID-19 patients, consistent with previous observations that anti-SARS-CoV-2 IgA is a marker of early SARS-CoV-2 infection^39-42^ and a potential driver of severe disease^41,43^ (**Figure 2A, D**). Conversely, overall CCP profiles were marked by enhanced antibody effector function despite having lower overall S-specific subclass titers, isotype titers, and FcR-binding. These observations suggest units of CCP, collected following the resolution of moderate SARS-CoV-2 infection, have qualitatively different antibodies with superior function. However, to fully capture the differences between CCP and recipient antibody profiles, a partial least square discriminant analysis (PLS-DA) was utilized to identify the antibody features that most differed across the antibody profiles (**Figure 2B, Supplemental Figure 2B**). Significant separation was observed across the SARS-CoV-2 antibody profiles. Among the top features that drove separation, 6 features were enriched in CCP and 4 features were enriched in the severe COVID-19 patients. CCP selectively exhibited enhanced levels of multiple measures of antibody function including antibody-dependent cell phagocytosis (ADCP), antibody-dependent NK cell activation (ADNK), and antibody-dependent complement deposition (ADCD)(**Figure 2C**) that were differentially enhanced across the serum samples (**Supplemental Figure 2C**). Conversely, IgM titers and IgA-binding FCAR responses were selectively expanded in severely ill CCP recipients. To further define the overall changes in the humoral immune response across the two groups, a correlation network was constructed with the additional humoral features that were significantly associated with the top 14 features (**Figure 2D**). Two separate networks appeared. The larger network including all the CCP enriched functional features and surprisingly the severe COVID-19 patient enriched RBD-specific IgM titer as well. FcR binding and IgG3 levels linked the CCP enriched features with RBD IgM. The tight correlation of S-specific ADCD and RBD IgM at the center of this network, suggests this profile of complement-inducing IgM antibodies is present in both groups. Given the profound lymphopenia seen in severe COVID-19,^5^ persistence of IgM responses in CCP units, a T-independent B1 cell response^44^ may be driving the IgM-enriched response shared between convalescents and severe COVID-19 patients. The smaller network of largely IgA and FCAR features also was primarily enriched in severe COVID-19 patients (**Figure 2D**). Thus, not unexpectedly, significant differences exist across CCP and severe COVID-19 patient plasma samples, with an enrichment of highly functional FcR-binding IgG3 SARS-CoV-2 specific antibodies in CCP and expanded IgM and IgA-centric responses in severely ill individuals, possibly related to ongoing viral replication within their respiratory tract.

**Figure 2.**
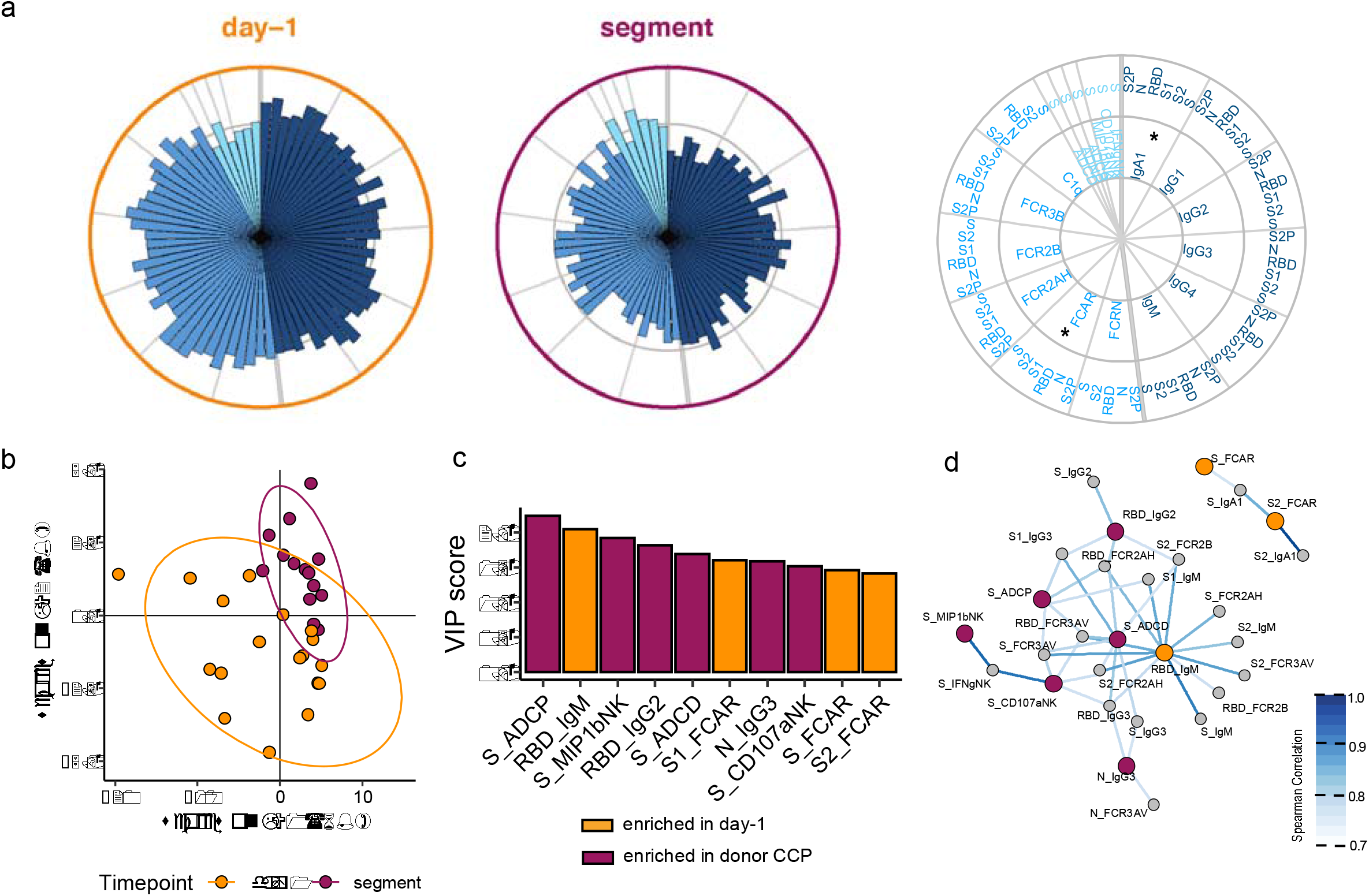
Functional Antibodies are Enriched in Convalescent Plasma. Severe COVID-19 patients and matched donor CCP were profiled for SARS-CoV-2-specific antibody responses. A) The polar plots depict the mean percentiles of SARS-CoV-2-specific antibody features within the day - 1 (n=18) and donor CCP (n=14) groups for the antigens S2P, N, RBD, S1, S2, and S. Each wedge represents a SARS-CoV-2 antibody feature, and the size of the wedge indicates the magnitude of the value. The colors represent the feature group: dark blue-antibody isotypes and subclasses; blue – Fc-receptor binding levels; light blue – antibody-dependent functions. Asterisks indicate global p-values obtained by non-parametric combination (Mann-Whitney U test p-values for partial tests within each feature type, and Fisher’s method for combination, *p < 0.05). Partial least squares discriminant analysis (PLS-DA) was applied to identify features distinguishing convalescent from day −1 severe COVID-19 patients. B) PLS-DA scores plot for the first two latent variables. Each dot is one sample, and the ellipses indicate 95% confidence regions assuming a multivariate t distribution. The model achieved an average cross-validation accuracy of 0.72. C) Variable importance in projection (VIP) score plot showing the top 10 important antibody features out of the 81 antibody features used to construct the PLS-DA. The color of the bar indicates in which group the feature is enriched, i.e., has a higher median value. D) Co-correlation network of features from the PLS-DA model in B with the top 10 VIP scores. The color of the nodes indicates whether the feature was enriched in donor CCP vs. day-1 sever COVID-19 patients. Nodes that were not top features were colored grey. Nodes enriched in day-1 were colored in orange and nodes enriched in donor CCP were colored in maroon. Edges were included if Spearman correlation was statistically significant after Benjamini-Hochburg correction of p values and the value of coefficient R was above 0.75 and the strength of the correlation was represented by the color of the edge.

### SARS-CoV-2-specific Antibody Profiles Globally Increase in Severe COVID-19 CCP Recipients

Despite the significant differences in antibody profiles across severe COVID-19 patient plasma and CCP, it is unclear whether the administration of CCP influences the overall evolution of the humoral immune response in the recipient patients. Thus, we began to look at the trajectory of humoral evolution across the severe COVID-19 patients in this clinical cohort (**Figure 3A**). For example, the majority of patients experienced a rapid and robust increase of IgG1-S-specific titers over time (**Figure 3A**). Similarly, an explosion of S-specific subclass, isotype, FcR binding, and functional responses were observed as early as day1 and later at day3 of the study follow-up (**Figure 3B** and **Supplemental Figure 3A**). Moreover, to gain specific insights into the features that evolved most substantially over time, we used multivariate models to compare antibody profiles across the cohort at pre-CCP and day 1 after CCP treatment (**Figure 3C**) as well as pre-CCP from day 3 after CCP treatment (**Supplemental Figure 3D**). Significant differences were noted in the multivariate models (**Supplemental Figure 3E**), marked by a largely expanding humoral immune response, with the evolution of less functional antibody subclasses including IgG4 and IgG2 responses to several antigens including to the RBD, S, S1. Conversely, two features were lost over time, including S1-specific FCAR binding and N-specific antibody binding to the neonatal FcR (FcRn) **(Supplemental 3B**,**C)**. However, finally, we examined the relationship between CCP antibody functions and the evolving humoral immune response in the severely ill COVID-19 patients in our patient cohort (**Figure 3E**). S-specific NK cell chemokine (MIP1β) and degranulation (CD107a) exhibited a trend towards a positive correlation with the evolving S-specific response in the COVID-19 patients, CCP-derived S-specific ADCP tended to negative correlation with the evolution of several FcR-binding, complement binding, ADCP, ADCD, and ADNP binding antibodies. Similarly, S-specific ADCD in CCP units was weakly negatively correlated with nearly all antibody features, except S-specific IgG4 levels. These data suggest that CCP-derived complement fixing antibodies could modulate the evolution of S-specific inflammatory humoral immunity in severe COVID-19 infection.

**Figure 3.**
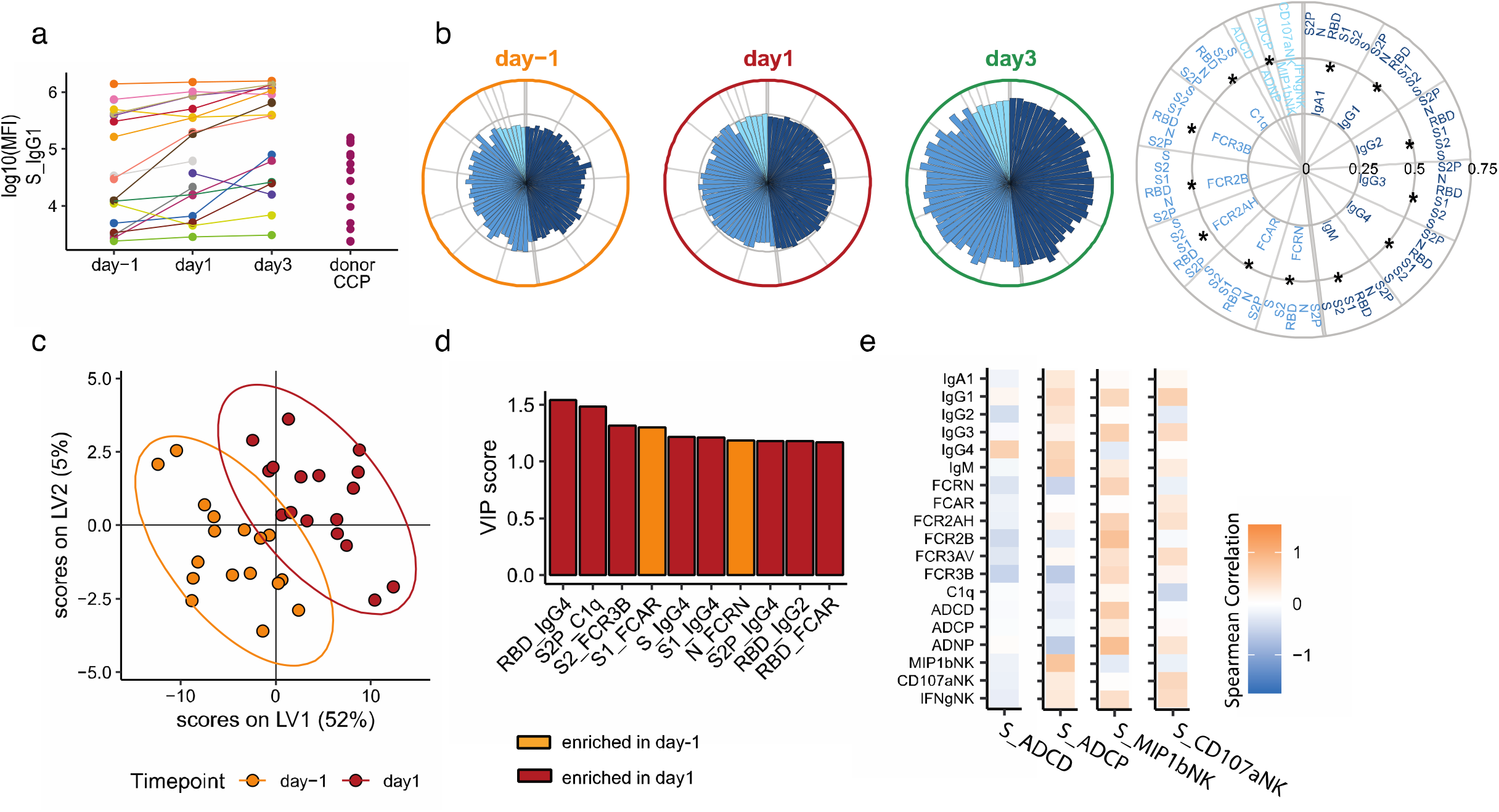
SARS-CoV-2-Specific Antibody Profiles Globally Increase in Severe COVID-19 CCP Recipients. Severe COVID-19 patients were profiled for SARS-CoV-2-specific antibody responses from day before receiving CCP (day −1) to one and three days after receiving CCP. **A)** SARS-CoV-2 S-specific IgG1 titers of COVID-19 patients at day −1 (n=18), day 1 (n=18) and day 3 (n=16), and 14 donor CCPs. Values are reported as log10 MFI. **B)** The polar plots depict the mean percentiles of SARS-CoV-2-specific antibody features within the day - 1 (n=18), day 1 (n=18) and day 3 (n=16) groups for the antigens S2P, N, RBD, S1, S2, and S. Each wedge represents a SARS-CoV-2 antibody feature, and the size of the wedge indicates the magnitude of the value. The colors represent the feature group: dark blue - antibody isotypes and subclasses; blue – Fc-receptor binding levels; light blue – antibody functions. An asterisk indicates global p-values obtained by non-parametric combination (Friedman test p-values for partial tests within each feature type, and Fisher’s method for combination, *p < 0.05). **C)** Multi-level partial least squares discriminant analysis (mPLS-DA) scores plot for the first two latent variables. Each dot is one sample, and the ellipses indicate 95% confidence regions assuming a multivariate t distribution. Colors indicate the time point the samples of the n=17 individual patients were taken (day −1 and day 1). The model achieved an average cross-validation accuracy of 86%. **D)** Variable importance in projection (VIP) score plot showing the top 10 important antibody features out of the 81 antibody features used to construct the mPLS-DA. The color of the bar indicates in which group the feature is enriched, i.e., has a higher median value. **E)** Heatmap showing the Spearman correlation coefficients between increases in antibody levels between day −1 and day 1 (y-axis) and corresponding donor CCP Spike-specific antibody features enriched in CCP including ADCD, ADCP, NK expression of MIP1b and NK expression of CD107a.

### Pre-existing Spike IgG1 Titer Determines the Effect of CCP on Severe COVID-19 Patients

Clinical experience suggests that a patient’s pre-existing antibodies affect the therapeutic benefit of transferred anti-Spike monoclonal antibodies^45^. Initial clinical studies of CCP in immunocompromised patients suggest that CCP can act a replacement for an absent humoral anti-SARS-CoV-2 response^46-48^. Whether this limited therapeutic window relates to the inability of antibody therapeutics to compete with pre-existing antibodies and function as a replacement for patient’s endogenous humoral response or is related to the need for distinct antibody functions in the setting of pre-existing immune complexes remains unclear. Thus, we split the severe COVID-19 patients into those with pre-transfusion high or low S IgG titers (**Figure 4A**). SARS-CoV-2 antibody evolution was observed in both the low and high pre-existing S-IgG cohorts (**Figure 4A**). To begin to define the role of CCP on shaping the evolution of the humoral immune response, we compared CCP antibody profiles to the change in antibody features in the severe COVID-19 patients (day-1 **→** 1) over time in the high and low pre-existing S IgG groups (**Figure 4B and C**). Unexpectedly, CCP profiles had distinct effects on shaping the evolution of the humoral immune response in the COVID-19 patients depending on their pre-existing S-specific titers. Specifically, while all antibody features were largely positively correlated across CCP and COVID-19 patients, N-specific humoral immune responses in the units of CCP were largely negatively correlated with all SARS-CoV-2 specific evolutionary profiles in the high pre-existing S-specific IgG titer group (**Figure 4B**). Conversely, more variable relationships were noted between CCP profiles and SARS-CoV-2 specific immune evolution in individuals with low pre-existing SARS-CoV-2 titers **(Figure 4B and C)**. Overall analyses of CCP features that modulated COVID-19 antibody profiles, across all subjects, pointed to a potential role for S-specific ADNP, S1-IgG3, RBD-C1q, N-IgA1, and N-IgG3 associated with significant changes in day-1**→**1 COVID-19 patient profiles depending on pre-existing S-IgG1 titer (**Figure 4D**). Further, deeper analysis of CCP features that were associated with day-1**→**1 COVID-19 patient humoral evolution across individuals that had high or low IgG titers at the start of therapy exhibited unique relationships. Specifically, S1-IgG3 and N-IgG3 were associated with attenuated evolution of inflammatory antibody profiles only among individuals that began with high titers on day1 **(Figure 4E)** and day 3 after CCP **(Supplemental Figure 5F)**, pointing to a potential role for highly functional antibodies in limiting inflammatory antibody evolution. Conversely, S-ADNP was highly associated with limiting inflammatory antibody evolution only in individuals that initially had low S-IgG **(Figure 4F)** titers pointing to a role for neutrophil mediated clearance by S-specific humoral immune responses as a potential mechanism for limiting the evolution of the inflammatory humoral immune response among individuals that began with low IgG titers. These data suggest that distinct populations of antibodies have immunomodulatory effects in patients with different levels of pre-existing IgG1 S-antibodies, with diverging roles for N- and S-specific responses in modulating responses at distinct starting levels of S-specific immunity. Thus, collectively these data highlight the unexpected and differential effect of CCP on modulating the evolution of SARS-CoV-2 immunity, dependent on pre-existing antibody levels.

**Figure 4.**
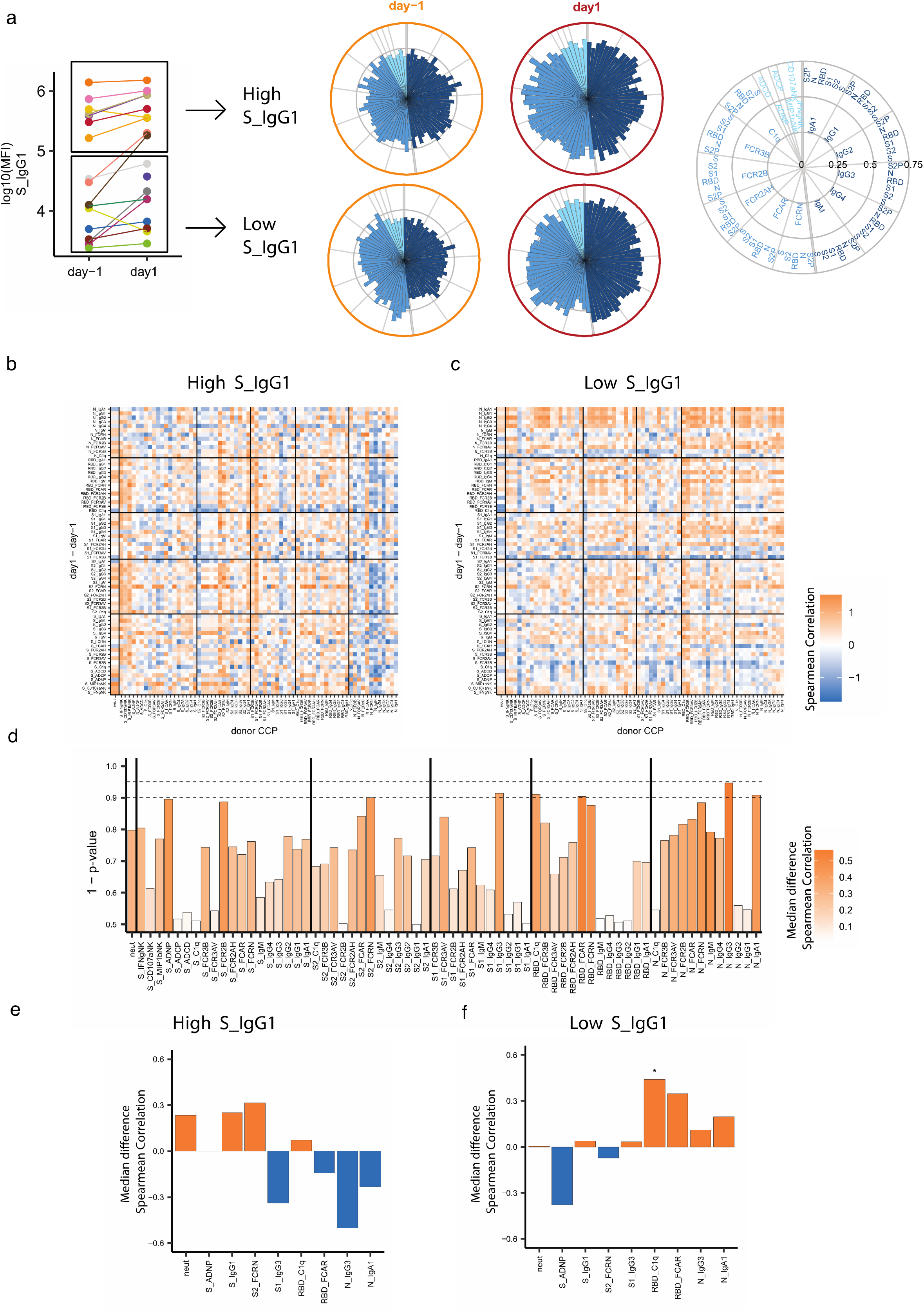
Effect of Convalescent Plasma on Recipient Antibody Trajectory is Influenced by Patient Pre-existing Spike IgG1 Titers. The effect of CCP antibody features on the trajectory (day 1 vs day-1) of the SARS-CoV-2 humoral response was profiled separately for severe COVID-19 patients with high and low pre-existing S IgG1 antibodies. **A)** SARS-CoV-2 S-specific IgG1 titers of COVID-19 patients at day −1 (n=18), day 1 (n=18). Patients were separated by the mean day-1 Spike IgG1 titer. Values are reported as log10 MFI. The polar plots depict the mean percentiles of SARS-CoV-2-specific antibody features within the day −1 (n=18), day 1 (n=18) groups for the antigens S2P, N, RBD, S1, S2, and S in the High and Low Spike IgG1 groups. Each wedge represents a SARS-CoV-2 antibody feature, and the size of the wedge indicates the magnitude of the value. The colors represent the feature group: dark blue - antibody isotypes and subclasses; blue – Fc-receptor binding levels; light blue – antibody functions. An asterisk indicates a global p-values obtained by non-parametric combination (Friedman test p-values for partial tests within each feature type, and Fisher’s method for combination, *p < 0.05). **B**,**C)** Heatmap showing the Spearman correlation coefficients between increases in antibody levels between day −1 and day 1 (y-axis) and corresponding donor CCP Spike-specific antibody features (x-axis) in patients with high (B) and low (C) levels of pre-existing Spike IgG1 antibody. **D)** Bar plot representing the statistical significance by permutation testing of the median difference between CCP features and patient trajectories in High and Low pre-existing IgG1 Spike patients. The Color represents the absolute value of the median difference and the height of the bar represents the p-value determined by the permutations test. **E**,**F)** Bar plot of the feature in (D) with P value <= 0.1 and Spike IgG1 and Neutralizing antibody titer. The color of the bars represents the median spearman correlation of CCP features in high pre-existing Spike IgG1 (E) and low pre-existing Spike IgG1 (F). An asterisk represents a CCP donor antibody feature with global p-values obtained by non-parametric combination (permutation test p-values for partial tests within each paired feature, and Fisher’s method for combination, *p < 0.05) within high (E) and low (F) preexisting Spike IgG1 patients.

## Discussion

Early in the pandemic, the absence of therapeutics, monoclonal antibodies, and vaccines sent the global biomedical community on an urgent hunt for potential treatments for severe COVID-19. Among the early therapeutics, CCP was proposed as a possible therapeutic due to its long successful history in the treatment of many diseases^49^, however the efficacy of CCP has been variable in COVID-19 viral pneumonia^38^. Why benefit is observed in some but not all studies is incompletely understood. Among the potential underlying reasons for this observed variable immune benefit may be differences in the quality of the CCP product. Importantly, beyond SARS-CoV-2-specific IgG titers and neutralization, antibodies contribute to antiviral immunity via a multitude of mechanisms that extend beyond binding and neutralization, mediated in part by the Fc-domain^50-52^. However, whether CCP units differ by Fc-characteristics and whether these qualitative differences influence patient plasma profiles was largely unknown. Here we observed striking heterogeneity in CCP Fc-profiles, enriched for functional antibodies, that influenced humoral evolution in severe COVID-19 recipients.CCP effects varied by pre-existing SARS-CoV-2 specific IgG titers at the time of therapeutic administration, revealing unexpected antigen-specific immunomodulatory effects of CCP.

Based on historical studies and vaccine trials across pathogens, pathogen-neutralization has been regarded as the key mechanism by which antibodies may confer protection and therapeutic benefit^35,53^. Along these lines, mounting evidence from non-human primate vaccine studies^54^ and epidemiologic studies^55^ of COVID-19, have demonstrated the association of neutralizing antibodies with prophylactic protection. Following suit, a recent Dutch clinical trial of CCP was discontinued early, despite being on pace to meet enrollment, because the majority of the patient population already had detectable neutralizing Ab titers at the time of potential CCP administration^24^. However, neutralization has not be linked to natural resolution of infection^30^, and instead has been linked to disease progression^56^. Further, the therapeutic benefit of IVIG, a pool of non-pathogen specific antibodies, in COVID-19^57^ has raised the possibility that antibodies may provide protection via alternative mechanisms, non-neutralizing and even non-pathogen specific mechanisms, such as reducing inflammation via the ligation of FcγR IIb or DC-SIGN^58^. To this point, emerging data suggests CCP can mediate immunomodulation, measured by a decrease in serum cytokines, in severe COVID-19 ^59^. Alternatively, antibodies could simply drive rapid viral clearance of pathogens or infected cells via complement activation, cytotoxic destruction, or opsonization of the virus or infected cells^50^. In a polyclonal therapy like CCP with multiple populations of antibodies, anti-inflammatory and pro-inflammatory antibodies aren’t mutually exclusive. Thus, a broader view of CCP functionality may provide us with clues to optimize this therapy for SARS-CoV-2 and for future pandemics.

Unlike any other COVID-19 therapeutics, CPP offers the twofold advantage of potentially modulating the inflammatory response while also providing anti-SARS-CoV-2 immunity. Comparison of CCP units demonstrated surprising heterogeneity, with variable patterns of isotype, subclass, and Fc receptor binding. However, despite this heterogeneity, CPP units exhibited more highly functional antibodies than severe COVID-19 patients. Differences between acute severe COVID-19 infection and convalescence are not surprising, given the dramatic expansion of humoral immune responses in early disease, particularly in those with more severe symptoms captured in this cohort^60^. Nevertheless, the data suggest that the expansion of overall humoral immune responses is not proportional to the functional evolution of the humoral immune response. Specifically, higher antibody titers were noted in severe COVID-19 patients that continued to increase over the course of illness. However, units of CCP had lower S-specific antibody titers, either due to the fact that they experienced less severe disease or due to the less inflamed nature of their immune response that persists for weeks to months after infection ^61-63^. Critically, relationships were observed between CPP antibody profiles and the evolution of the severe COVID-19 profiles, that were negligibly driven by SARS-CoV-2 S titers or neutralization, but interesting driven by particular SARS-CoV-2-specific subclass levels and functions.

Two recent studies of anti-RBD monoclonal antibodies have shown that Fc effector functions are essential for the therapeutic benefit in mice and hamster models of COVID-19^64,65^. Likewise, here we show that specific antibody effector functions may represent key functional mechanisms critical for antiviral immunity. S-specific ADNP activity was anticorrelated with the SARS-CoV-2 antibody trajectory. Though typically thought of as responsive to primarily bacterial infections, animal models of poxviruses and influenza A have shown that neutrophils are the first responders in the immune system to respiratory viruses^66,67^. Our data suggests that enhancing the phagocytic capabilities of the first responding cells, possibly by enhancing viral clearance before subsequent infiltration of monocytes implicated in COVID-19 hyperinflammation^68-70^, may play a critical part in blocking worsening inflammation. Further S-specific ADNP was anti-inflammatory only in patients with low-pre-existing antibody. Since IgG S titer is a surrogate for disease progression, these data suggest that enhanced neutrophil phagocytosis can preferentially modulate the inflammatory cascade early on in disease before subsequent immune actors hone to virus-infected parts of the respiratory tract. Potentially, antibody enhancement of neutrophil phagocytosis may be an important part of the success seen in early administration of CCP to outpatients^19^, preventing the transition of mild controlled viral infection to unchecked inflammation of severe COVID-19. Given the use of highly analogous Fc-receptors, FCGR3a and FCGR3b, on NK cells and neutrophils, respectively, it is possible that efforts to enrich for ADCC may however inadvertently recruit CCPs able to also co-recruit neutrophil function. While ADCC was not associated with differential profiles in our patient population, pointing to neutrophils as key responders, it remains possible that neutrophils and NK cells may function together to clear and dampen infection. However, in our efforts to enrich for neutralization, we may be missing the selection of units of CCP able to strongly recruit ADNP effector function and stacking the deck against ourselves.

Even if we identify the most potent CCP, a body of literature suggest it may not be effective if given too late in COVID-19 disease. Focusing on randomized control trials, CCP resulted in a 73% risk reduction COVID-19 progression when given within 3 days of symptoms^19^, but has not had a mortality benefit in randomized clinical trials when given latter in illness to severe COVID-19 patients^20^. Similarly, for monoclonal antibody therapies, Phase I-III clinical trials of Regeneron’s cocktail of two anti-RBD monoclonals (REGN-COV2) demonstrated a greater improvement on viral load and progression of COVID-19 in outpatients without detectable antibodies^45^. Importantly in order to benefit the patient, CCP transferred antibodies have to compete with existing antibodies to traffic to foci of infection, displace existing antibodies within immune complexes, and compete with existing antibodies for Fc-receptors and lectins on immune cells. In patients with high pre-existing S-specific antibodies, this competition may be insurmountable for the numbers of transferred antibodies with 1-2 units of CCP. However, if functionally optimized, even small numbers of antibodies may be sufficient to tip the balance of pre-existing antibody pools. Quite interestingly, in individuals with high pre-existing S antibody levels we found multiple N-specific antibody features in CCP tracked with dampened inflammatory evolution of SARS-CoV-2 specific humoral immunity in patients. Thus, the displacement or replacement of N-specific antibodies with antibodies from CCP appeared to have an unexpected therapeutic benefit. Nucleocapsid is a viral RNA-binding multifunctional protein highly expressed in corona virus-infected cells^71,72^. Antibody responses to this immunodominant antigen can precede S-specific antibodies in some patients^73^ but also may be associated with progression to severe disease when involved in robust complement fixation ^30^. Thus, the data presented here point to a potential role of less-inflammatory CCP derived N-specific antibodies as a potential driver of less inflammatory disease. N-specific-CCP-derived antibodies could displace more inflammatory patient-derived antibodies that may contribute to enhanced pathology rather than immune protection. Altogether, the interplay of CCP antibody profiles and sever COVID-19 patients has demonstrated the critical role of pre-existing humoral immunity in the potential immunomodulatory effects of CCP via both S- and N-specific antibodies.

This study of patients enrolled in Expanded Access Program early on in the COVID-19 pandemic was unable to address questions related to mortality or to control for the natural evolution of the humoral immune response but pointed to several unexpected mechanisms by which CCP may modulate humoral evolution in severe COVID-19 infection. Despite the limited sample small sample size of this cohort, we have been able to make fundamental observations about the heterogeneity of convalescent responses, functional antibodies that distinguish CCP from acute patient serum, and that the effect of CCP depends on patient’s pre-existing humoral immune response. The extension of these novel findings to larger, randomized control studies, will be essential to validate our findings. Altogether, we hope these mechanistic insights int CCP’s therapeutic benefit will provide unique hints for the rational design of next generation monoclonal therapeutics with a longer-window of therapeutic efficacy, even pointing to potential benefit of including N-specific therapeutics. Moreover, these data may even provide insights for how we collect, enrich, and use CCP in the future. Though CCP was thought of a therapy of necessity in March 2020 that would be replaced by more refined monoclonals, emerging COVID-19 variants are creating new therapeutic gaps that an improved CCP may be needed to fill. Collectively, this deeper look at CCP therapy provides a first glimpse at the unique opportunities to leverage CCP as a COVID-19 therapeutic and provides a potential future rationale for stratification of CCP donors for COVID-19 and future pandemics.

## Methods

### Antibody Titer and Fc-Receptor Binding Assays

Antigen-specific antibody subclass, isotype, and Fc-receptor (FcR) binding levels were assayed with a customized multiplexed Luminex bead array, as previously described^74^. This allows for relative quantification of antigen-specific humoral responses in a high-throughput manner and simultaneous detection of many antigens. A panel of SARS-CoV-2 antigens including the full spike glycoprotein (S) (provided by Lake Pharma), receptor binding domain (RBD) (Provided by Aaron Schmidt, Ragon Institute) nucleocapsid (N) (Aalto Bio Reagents, Dublin, Ireland), S1 (Sino Biological, Beijing, China) and S2 (Sino Biological, Beijing, China) were used. Control antigens were run including a mix of three Flu-HA proteins (H1N1/A/New Caledonia/20/99, H1N1/A/Solomon Islands/3/2006, H3N2)(A/Brisbane/10/2007 - Immune Tech) and Ebola glycoprotein (IBT Bioservices). In brief, antigens were coupled to uniquely fluorescent magnetic carboxyl-modified microspheres (Luminex Corporation, Austin, TX) using 1-Ethyl-3-(3-dimethylaminopropyl) carbodiimide (EDC) (Thermo Fisher Scientific, Waltham, MA) and Sulfo-N-hydroxysuccinimide (NHS) (Thermo Fisher Scientific, Waltham, MA). Antigen-coupled microspheres were then blocked, washed, and incubated for 16 hours at 4°C while rocking at 700 rpm with diluted plasma samples at plate concentrations of 1:12,000 for all subclasses and isotypes and C1q and Fcrn binding and 1:120,000 for all other Fc-receptors to form immune complexes in a 20 uL volume in 384 well plates (Greiner, Monroe, NC). The following day, plates were washed using an automated plate washer (Tecan, Männedorf, Zürich, Switzerland) with 0.1% BSA and 0.02% Tween-20. Antigen-specific antibody titers were detected with Phycoerythrin (PE)-coupled antibodies against IgG1, IgG2, IgG3, IgG4, IgA1, and IgM (SouthernBiotech, Birmingham, AL). To measure antigen-specific Fc-receptor binding, biotinylated Fc-receptors (FcR2AH, 2B, 3AV, 3B, FCRN, FCAR, FCR3AV - Duke Protein Production facility, C1q – Sigma Aldrich) were coupled to PE to form tetramers and then added to immune-complexed beads to incubate for 1 hour at room temperature while shaking. Fluorescence was detected using an Intellicyt iQue with a 384-well handling robot (PAA) and analyzed using Forecyt software by gating on fluorescent bead regions and PE mean fluorescence intensity (MFI) was measured as the readout of each antigen-specific antibody measurements. All experiments were performed in duplicate while operators were blinded to study group assignment and all cases and controls were run at the same time to avoid batch effects.

### Ab-Directed *Functional Assays*

Bead-based assays were used to quantify antibody-dependent cellular phagocytosis (ADCP), antibody-dependent neutrophil phagocytosis (ADNP) and antibody-dependent complement deposition (ADCD) in the MGH SARS-CoV-2 cohort, as previously described^75-79^. Yellow (ADNP and ADCP) as well as red (ADCD) fluorescent neutravidin beads (Thermo Fisher) were coupled to biotinylated SARS-CoV-2 S antigens and incubated with diluted plasma (ADCP 1:100, ADNP 1:50, ADCD 1:10) to allow immune complex formation for 2h at 37°C. To assess the ability of sample antibodies to induce monocyte phagocytosis, THP-1s (ATCC) were added to the immune complexes at 1.25E5cells/ml and incubated for 16h at 37°C. For ADNP, primary neutrophils were isolated via negative selection (Stemcell) from whole blood. Isolated neutrophils at a concentration of 50,000 per well were incubated with immune complexes for 1h incubation at 37°C. Neutrophils were stained with an anti-CD66b PacBlue detection antibody (Biolegend) and fixed with 4% paraformaldehyde (Alfa Aesar). To measure antibody-dependent deposition of C3, lyophilized guinea pig complement (Cedarlane) was reconstituted according to manufacturer’s instructions and diluted in gelatin veronal buffer with calcium and magnesium (GBV++) (Boston BioProducts) and mixed with immune complexes. After a 20-minute incubation at 37C, C3 was detected with an anti-C3 fluorescein-conjugated goat IgG fraction detection antibody (Mpbio). Antibody-dependent NK cell activity was measured via an ELISA-based assay, as described previously (Chung et al., 2015). Briefly, plates were coated with 3mg/mL of antigen (SARS-CoV-2 S) and blocked overnight at 4°C. NK cells were isolated the day of the assay via RosetteSep (Stem Cell Technologies) from healthy buffy coats (MGH blood donor center). Diluted plasma samples were added to the antigen-coated plates (1:25 dilution) and incubated for 2h at 37°C. NK cells were mixed with a staining cocktail containing anti-CD107a BV605 antibody (Biolegand), Golgi stop (BD Biosciences) and Brefeldin A (BFA, Sigma Aldrich). 2.5E5 cells/ml were added per well it the immune complexes and incubated for 5h at 37°C. Following, cells were fixed (Perm A, Invitrogen) and stained for surface markers with anti-CD3 APC-Cy7 (BioLegend) and anti-CD56 PE-Cy7 (BD Biosciences). Subsequently, cells were permeabilized using Perm B (Invitrogen) and intracellularly stained with an anti-MIP1b-BV421 (BD Biosciences) and IFNg-PE (BioLegend) antibodies.

All assays were acquired via flow cytometry with iQue (Intellicyt) and an S-Lab robot (PAA). For ADCP, events were gated on singlets and bead-positive cells. For ADNP, neutrophils were defined as CD66b positive events followed by gating on bead-positive neutrophils. A phagocytosis score was calculated for ADCP and ADNP as (percentage of bead-positive cells) x (MFI of bead-positive cells) divided by 10000. ADCD was reported as the median of C3 deposition. NK cells were defined as CD3- and CD56+ events. Data were reported as the percentage of cells positive for CD107a, MIP-1β, and IFNg.

### rVSV-SARS-CoV-2 S Neutralization Assay

The neutralization assay was done as previously described ^80^. Briefly, CCP samples were serially diluted and incubated with pre-titrated amounts of virus for 1 hr at RT, plasma-virus mixtures were added to 96-wellplates (Corning) containing monolayers of Vero cells, incubated for 7hr at 37°C/5% CO2, fixed with 4% paraformaldehyde (Sigma) in PBS, washed with PBS, and stored in PBS containing Hoechst-33342 (1:2,000 dilution; Invitrogen). Viral infectivity was measured by automated enumeration of green fluorescent protein (GFP)-positive cells from captured images using a Cytation5 automated fluorescence microscope (BioTek) and analyzed using the Gen5 data analysis software (BioTek). The serum half-maximal inhibitory concentration (IC50) was calculated using a nonlinear regression analysis with GraphPad Prism software.

### Data Pre-processing

Antibody isotypes, subclasses, FcR-binding levels and ADCD measurements were log10 transformed. Antibody features were excluded from the analysis if the maximal value across samples and time points (day −1, day 1, day 3 and donor CCP) was less than four standard deviations above the mean value obtained for PBS controls. Three features (N_FCR2AH, S1_FCRN, S1_C1q) were excluded.

### Polar Plots

Polar plots for Figures 2,3, 4 were used to visualize the mean percentile of groups. Percentile rank scores were determined for each feature across all considered samples using the function ‘percent_rank’ of the R package ‘dplyr’.

Polar plots for Figure 1 and Supplemental Figure 2 were used to visualize the S-specific individual antibody profile of CCP units and pre-CCP severe acute COVID-19 patients (day-1). Each feature across the respective populations were scaled by min-max normalization. The order of individual polar plot was determined as follows: first, sample-sample similarities of S-specific antibody profiles were calculated using the function ‘dist’ of the R package ‘stats’. Then, the samples were clustered to generate the dendrogram tree by the agglomeration clustering function ‘hclust’ with default parameters in the R package ‘stats’. The dendrogram tree was sorted using the function ‘dendsort’ of the R package ‘dendsort’ to optimize the ordering of samples. Finally, the polar plots were organized based on the order of sorted dendrogram tree.

### Non-parametric Combination

Global statistical differences of feature types (e.g. IgG1) across antigens and between groups were assessed using non-parametric combination^81,82^. For each feature type, partial test p-values were obtained by Mann-Whitney U test for the comparison of day −1 and segment, and Friedman tests for the comparison of day −1, day 1 and day 3 measurements for each antigen-specific sub-feature (e.g., S_IgG1, N_IgG). The partial p-values were combined using Fisher’s method to obtain a global statistic. This procedure to obtain a global statistic was repeated 1000 times for data with permuted group labels, preserving the permuted structure for the partial tests, and was used to construct a null distribution of global statistics. For the analysis of day −1, day 1 and day 3, only patients were included that had measurements at each time point, and for the permutations the labels were shuffled for each patient individually. The true global statistic obtained from the unpermuted data was compared to the null distribution and a global p-value was determined as the tail probability. For the functions which were only measured for SARS-CoV-2 S, the global p-value was obtained by merging all the functions.

### Testing for the Effect of CCP on Humoral Antibody Trajectories with Non-parametric Combination Testing of the Median Difference of Spearman Correlation

To evaluate the significance of correlation difference between high S_IgG1 and low S_IgG1 group, we permuted the group of recipient-donor pairs with the fixed proportion of high and low S_IgG1 group for 800 times and then calculated the median correlation of each feature with donor CCP features. After that, we estimated the p values as the proportion of permutated median correlation values of permutations above and below the observed the actual median. The features with a permutation test P value of correlation difference less than 0.1 along with S IgG1 and Neutralizing antibody titer were selected for further exploration.

To evaluate the effect of recipient-donor pair, global statistical differences of each feature in CCP across all the features in recipient were evaluated using non-parametric combination as described above. For high S_IgG1 or low S_IgG1 group respectively, we broke the recipient donor pair, randomly matched them, and calculated the Spearman correlation on the permuted recipient-donor pair 1000 times. The partial test p-values were obtained by spearman correlation and were combined using Fisher’s method to obtain a global statistic by comparing actual value from true recipient-donor pairs with the null distribution generated from permuted pairs for each feature in CCP.

### Multivariate Model

Partial least square discriminant analysis (PLS-DA) was performed to discriminate day −1 patient samples from donor CCP samples. Multi-level PLS-DA (mPLS-DA)^83^ was performed to discriminate day −1 and day 1/day 3 measurements and to take into account the paired structure of the data. For the multi-level PLS-DA, only patients were included that had measurements at both considered time points. Missing values were imputed using k-nearest neighbor imputation (R package ‘DMwR’) before z-scoring. For the mPLS-DA, the data was first imputed, denoised and then z-scored. The model performance was assessed in a five-fold cross-validation framework, and the average cross-validation accuracy was reported for 100 repetitions of cross-validation. For the mPLS-DA, the folds for cross-validation are generated such that measurements of the same patient are in the same fold. Variable importance of projection (VIP) scores, which describe the contribution of each feature to the model, were used to rank the features, and the top 10 important features were displayed. The modeling approach was validated using permutation tests. Control models with ‘permuted labels’ were generated, for which the model was trained and applied to data with shuffled group labels in the same cross-validation framework. For the case of paired data (each patient has measurements at both time points) in the mPLS-DAs, the labels were flipped with a 50% probability to obtain the control models. This procedure was done for 500 permutations for each of 10 cross-validation replicates. The p-values for the modeling approach were obtained from the tail probability of the generated null distribution, i.e., the distribution of classification accuracies of the control models. The median p-value across the 10 cross-validation replicates was reported in the figures. PLS-DA models were generated with the R package ‘ropls’ interfaced by R package ‘systemsseRology’ (https://github.com/LoosC/systemsseRology). The analyses were performed with R version 4.0.2.

### Correlation Analysis

We calculated Spearman correlation and their p-values using the R function ‘cor.test’ of the R package ‘stats’. After that, the p-values were adjusted by Benjamini-Hochburg procedure for multiple testing correction. The adjusted p-values were labeled by Asteriks (*: adjusted p value < 0.05) in the correlation heatmap if they were significant.

A chord diagram was used to visualize the significant links among the humoral features using the function ‘chordDiagram’ in R package ‘circlize’(0.4.12). To evaluate co-correlated relationships between top features selected by PLS-DA and additional humoral immune features, the significant Spearman correlations above a threshold of |r| > 0.75 were selected and a layout was created to specify the spatial position to maintain correlation patterns using the function ‘create_layout’ in R package ‘ggraph’, where the gradient color of links represented the values of correlations and the color of nodes denoted the enriched group. After that, the layout was visualized as the correlation network using the function ‘ggraph’ in the R package ‘ggraph’ (2.0.4). Additionally, the labels and positions of nodes and links were adjusted for better visualization using the software Adobe Illustrator 2020 (24.2.3).

## Data Availability

Data is available upon request

## Acknowledgements

We thank Nancy Zimmerman, Mark and Lisa Schwartz, an anonymous donor (financial support), Terry and Susan Ragon, and the SAMANA Kay MGH Research Scholars award for their support. We acknowledge support from the Ragon Institute of MGH, MIT and Harvard, the Massachusetts Consortium on Pathogen Readiness (MassCPR), the NIH (3R37AI080289-11S1, R01AI146785, R01-AI132633, U19AI42790-01, U19AI135995-02, U19AI42790-01, 1U01CA260476 – 01, CIVIC75N93019C00052, T32 AI007061), the Gates foundation Global Health Vaccine Accelerator Platform funding (OPP1146996 and INV-001650), and the Musk Foundation.

## Authorship Contributions

J.D.H., B.J., K.J.B., L.A.P, and G.A. conceived of the idea. H.Y., J.R., M.E.D., D.H., R.K.J.,R.H.B., K.C., and L.A.P. designed, conducted, analyzed the clinical cohort, and performed neutralization antibody titer measurements. J.D.H. and G.A. designed the experiments. J.D.H. performed the experiments except for the neutralization antibody titer measurements. J.D.H., C.W., and C.L. analyzed the data. And J.D.H., C.W., C.L., L.A.P, and G.A. wrote the paper with input from all authors.

