## Supplemental Table and Figures for "Functional Antibodies in COVID-19 Convalescent Plasma"

| Baseline Patient Characteristics | Mean | Median | | | | | |
|---|---|---|---|---|---|---|---|
| BMI | 27.1 | 27.2 | | | | | |
| Age | 63.2 | 61.0 | | | | | |
| Gender | **Female (%)** | **Male (%)** | | | | | |
| | 52.6 | 47.4 | | | | | |
| Ethnicity | **Hispanic (%)** | **Non-Hispanic (%)** | **Other/Not Reported (%)** | | | | |
| | 47.4 | 47.4 | 5.3 | | | | |
| Race | **White (%)** | **African American (%)** | **Asian (%)** | **Other/Not Reported (%)** | | | |
| | 10.5 | 26.3 | 10.5 | 52.6 | | | |
| Comorbidities | **Hypertension (%)** | **Diabetes (%)** | **Chronic Lung Disease (%)** | **Chronic Kidney Disease (%)** | **Coronary Artery Disease (%)** | **Heart Failure (%)** | **Hyperlipidemia (%)** |
| | 78.9 | 42.1 | 15.8 | 36.8 | 5.3 | 15.8 | 63.2 |
| **Clinical Characteristics on Enrollment in Study** | **Mean** | **Median** | | | | | |
| WHO Score - Day 0 | 6.2 | 5.0 | | | | | |
| O2 Status - Day 0 | **Room Air (%)** | **Nasal Cannula > 5L or non-rebreather (%)** | **Non-Invasive Ventilation, High-Flow Nasal Cannula (%)** | **Mechanical Ventilation (%)** | | | |
| | 0.0 | 52.6 | 21.1 | 26.3 | | | |
| | **Mean** | **Median** | | | | | |
| Day of Hospitalization CCP Administered | 1.7 | 1.0 | | | | | |
| **COVID-19 Treatments Administered During Study** | **Treated (%)** | **Not-Treated (%)** | | | | | |
| Corticosteroids | 94.7 | 5.3 | | | | | |
| Hydroxychloroquine | 47.4 | 52.6 | | | | | |
| Remdesivir | 5.3 | 94.7 | | | | | |
| Sarilumab | 5.3 | 94.7 | | | | | |
| Leronlimab | 5.3 | 94.7 | | | | | |
| **Clinical Outcomes of Participants** | | | | | | | |
| Clinical Status - Day 14 | **Discharged (%)** | **Hospitalized (%)** | **Deceased (%)** | | | | |
| | 57.9 | 21.1 | 21.1 | | | | |
| | **Mean** | **Median** | | | | | |
| WHO Scale - Day 14 | 4.4 | 3.0 | | | | | |
| Day of Discharge | 12.0 | 9.0 | | | | | |
| Mean Day of Death | 11.2 | 14.0 | | | | | |
| O2 Status - Day 14 (Survivors) | **Room Air (%)** | **Nasal Cannula > 5L or non-rebreather (%)** | **Non-Invasive Ventilation, High-Flow Nasal Cannula (%)** | **Mechanical Ventilation (%)** | | | |
| | 86.7 | 0.0 | 6.7 | 6.7 | | | |

**Supplemental Table 1.** Clinical Characteristics of CCP Recipients

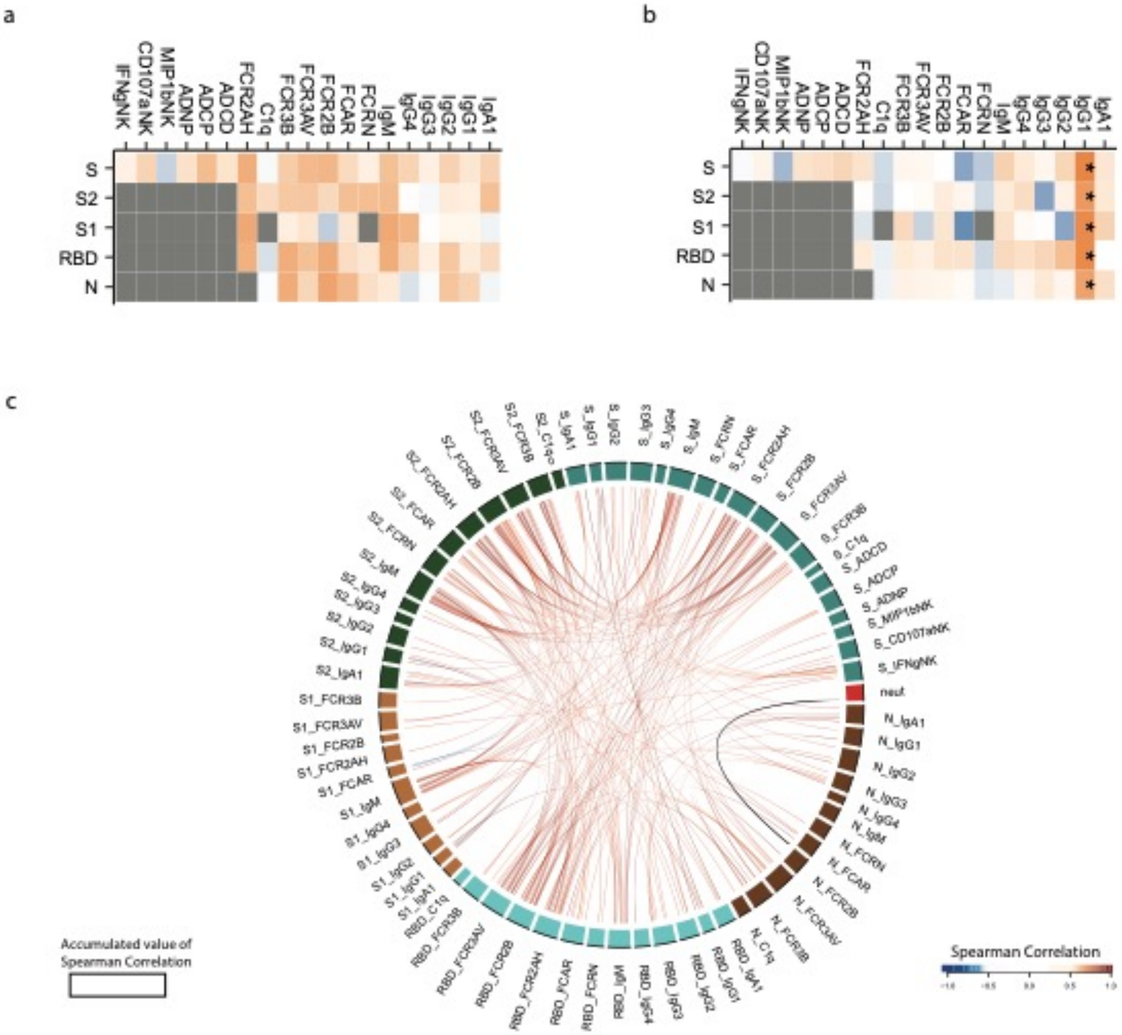


**Supplemental Figure 1.** Correlation of CCP Features

Correlation of Neutralizing antibody titer (A) and Spike IgG1 titer (B) with Spike, S1, S2, RBD, and Nucleocapsid-specific features. An asterisk represents a statistically significant correlation after multiple-test correction. Grey boxes represent assays that were not performed. (C) A cord diagram representing the Spearman correlation >0.75 among all SARS-CoV-2 antibody features in donor CCP. The strength of the correlation is represented by the color of the cord connecting the two nodes, with one exception, correlations with neutralizing antibody titer that are colored in black. The width of antibody each feature represents the accumulated values of Spearman correlation coefficients of that feature with all other features included in the diagram.

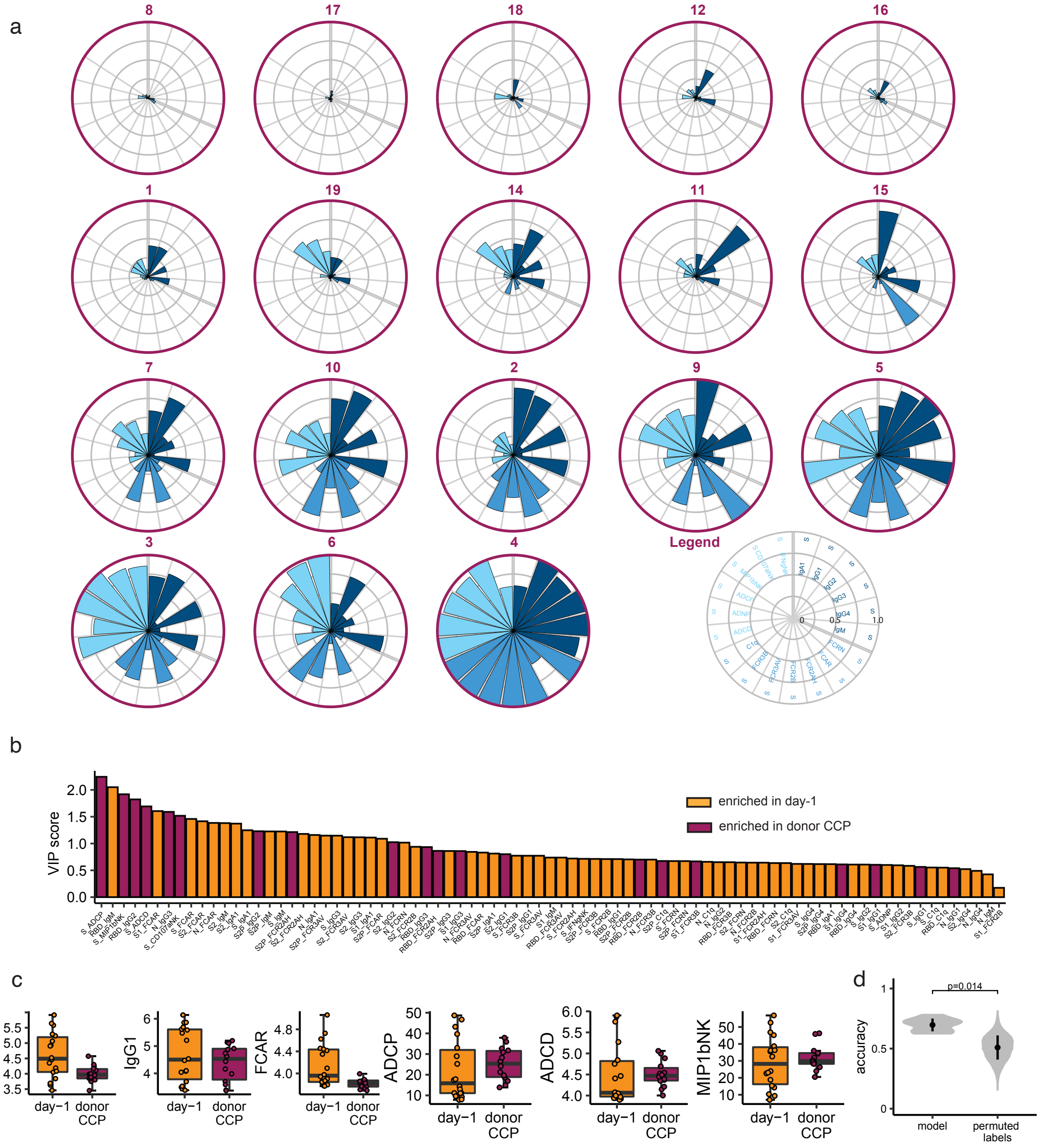

**Supplemental Figure 2.**

Supplemental figures supporting Figure 2, comparing the SARS-CoV-2 Specific antibody responses in CCP units vs. pre- CCP (day -1).

**A)** Day -1 severe COVID-19 patients were profiled for SARS-CoV-2 Spike-specific antibody responses. Each polar plot depicts an individual donor's anti-Spike antibody profile, scaled to the minimum and maximum of the 14 units of CCP profiled. Each wedge represents a SARS-CoV-2 antibody feature, and the size of the wedge indicates the magnitude of the value. The colors represent the feature group: dark blue - antibody isotypes and subclasses; blue – Fc-receptor binding levels; light blue – antibody-dependent functions; light grey - neutralizing antibody titer. Polar plots were organized by hierarchical clustering of S-specific antibody features. (B and C) Correlation matrix of neutralizing antibody titer.

**B)** Variable importance in projection (VIP) scores for all 81 antibody features. The color of the bar indicates in which group the feature is enriched, i.e., has a higher median value.
**C)** Boxplots showing examples of S-specific antibody features for patient samples at day -1 (n=18) and donor CCP (n=14). IgA1, IgG1, FCAR, and ADCD are reported as log10 MFI values, ADCP as phagocytosis score, and MIP1bNK as percentage of NK cells positive for MIP1b.
**D)** Accuracies from 10 repetitions of five-fold cross-validation for the actual model and models based on permuted labels. The permuted labels models are repeated 500 times for each cross-validation repetition. The p-values are determined based on the probability that the accuracy of the permuted label model is higher than for the actual model and is reported as median of 10 cross-validation repetitions.

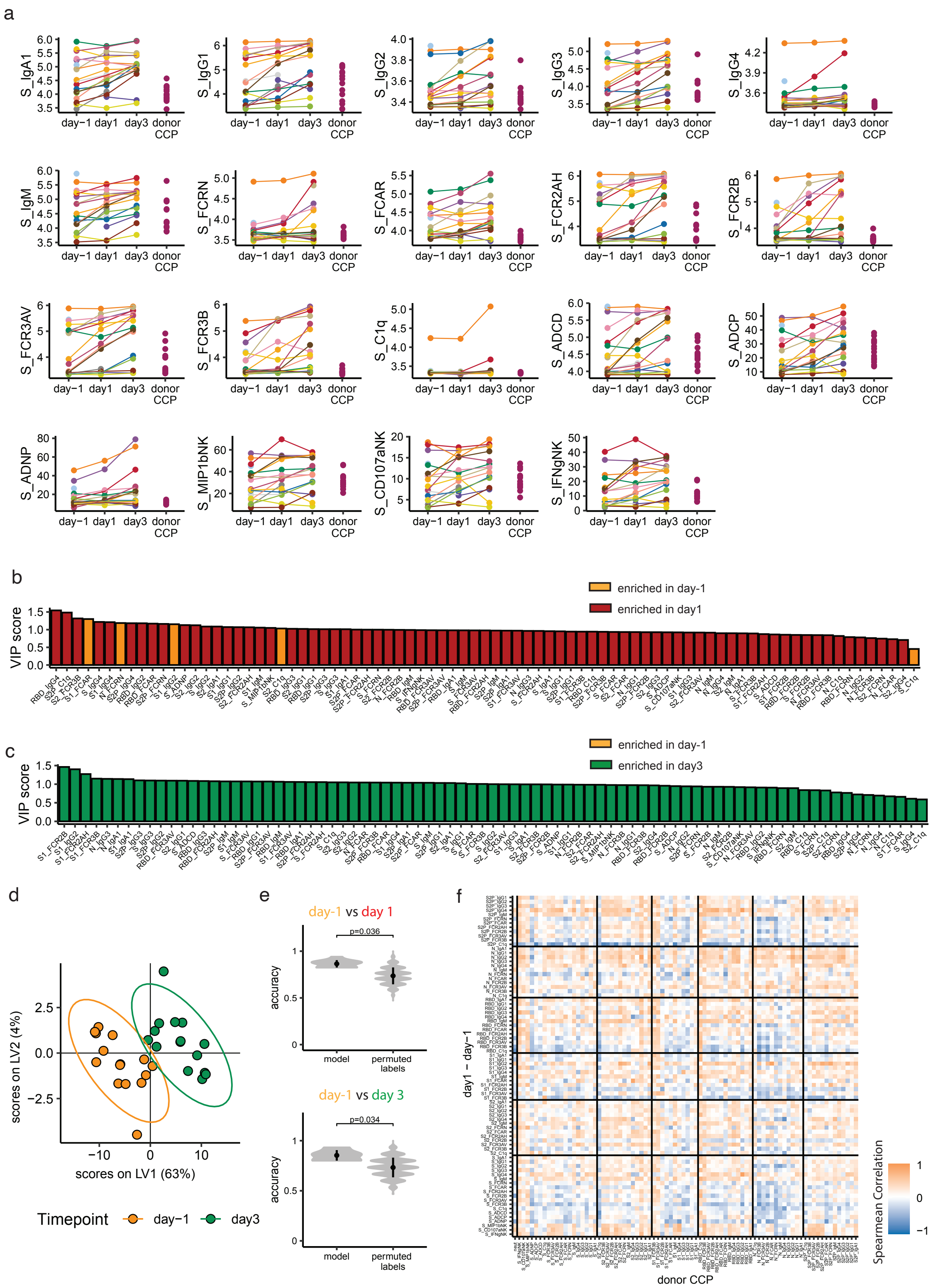

a
S_IgA1
S_IgG1
S_IgG2
S_IgG3
S_IgG4
S_IgM
S_FCRN
S_FCAR
S_FCR2AH
S_FCR2B
S_FCR3AV
S_FCR3B
S_C1q
S_ADCD
S_ADCP
S_ADNP
S_MIP1bNK
S_CD107aNK
S_IFNgNK
day−1 day1 day3 donor CCP
b
VIP score
enriched in day-1
enriched in day1
c
VIP score
enriched in day-1
enriched in day3
d
scores on LV2 (4%)
scores on LV1 (63%)
Timepoint
day−1
day3
e
day-1 vs day 1
p=0.036
accuracy
model
permuted labels
day-1 vs day 3
p=0.034
f
day1 − day−1
donor CCP
Spearman Correlation

**Supplemental Figure 3.**
Additional figures supporting Figure 3, demonstrating the difference in SARS-CoV-2 specific antibody responses between day -1 and CCP and day -3 and CCP.
**A)** SARS-CoV-2 S-specific antibody features for each of the 19 patients and 14 donor CCPs.
**B,C)** Variable importance in projection (VIP) score plot for the 81 antibody features used to construct the mPLS-DA. The color of the bar indicates in which group the feature is enriched, i.e., has a higher median value. (B) day -1 vs. day 1 mPLS-DA model. (C) day -1 vs. day 3 mPLS-DA model.
**D)** Multi-level partial least squares discriminant analysis (mPLS-DA) scores plot for the first two latent variables for the day -1 vs. day 3 mPLS-DA model. Each dot is one sample, and the ellipses indicate 95% confidence regions assuming a multivariate t distribution. Colors indicate the time point the samples were taken of the n=15 patients. The model achieved an average cross-validation accuracy of 86%.
**E)** Accuracies from 10 repetitions of five-fold cross-validation for the actual model and models based on permuted labels. The permuted labels models are repeated 500 times for each cross-validation repetition. The p-values are determined based on the probability that the accuracy of the permuted label model is higher than for the actual model and is reported as median of 10 cross-validation repetitions.
**F)** Heatmap showing the Spearman correlation coefficients between increases in antibody levels between day -1 and day 1 and corresponding donor CCP antibody levels.

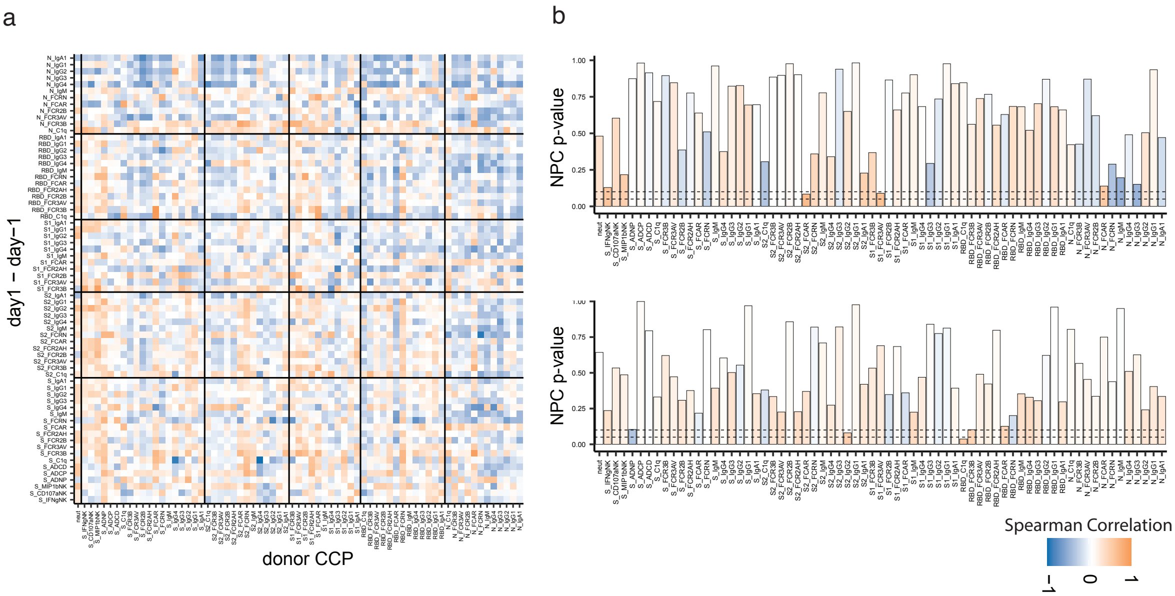


**Supplemental Figure 4.**

Additional figures supporting Figure 4, demonstrating the differences in the effect of CCP on evolution of the anti-SARS-CoV-2 humoral immune response depending on the pre-existing S IgG1 titers. (A): Heatmap depicting the difference of the Spearman correlation (donor CCP vs. day 1 – day-1) between the S_IgG1 high group and S_IgG1 low group. (B) Bar plots of Non-parametric Combination (NPC) test on each feature in donor CCP in High pre-existing S IgG1 (top) and Low pre-existing S IgG1 (bottom) individuals. Color in (A) represents the strength of the difference in correlation and in (B) represents the strength of the median Spearman correlation of each CCP feature in the respective High and Low pre-existing S IgG1 groups.

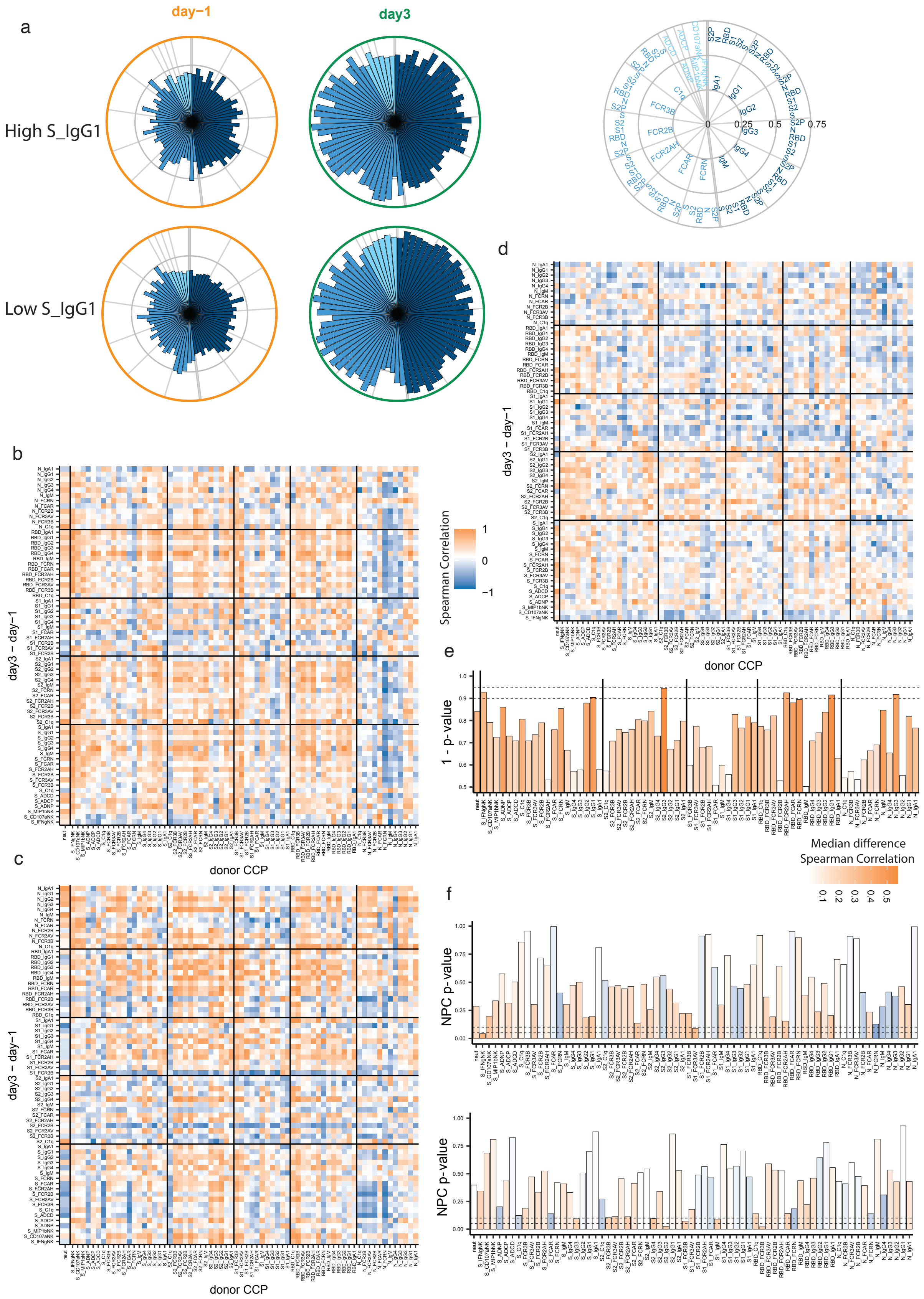
a
day−1
day3
High S_IgG1
Low S_IgG1
0
0.25
0.5
0.75
b
c
d
e
f
day3 − day−1
donor CCP
Spearman Correlation
1
0
−1
1 - p-value
Median difference
Spearman Correlation
NPC p-value

**Supplemental Figure 5.**
The effect of CCP antibody features on the trajectory (day 3 vs day-1) of the SARS-CoV-2 humoral response was profiles separately for severe COVID-19 patients with high and low pre-existing S IgG1 antibodies.
**A)** The polar plots depict the mean percentiles of SARS-CoV-2-specific antibody features within the day -1 (n=18), day 3 (n=16) groups for the antigens S2P, N, RBD, S1, S2, and S in the High and Low Spike IgG1 groups. Each wedge represents a SARS-CoV-2 antibody feature, and the size of the wedge indicates the magnitude of the value. The colors represent the feature group: dark blue - antibody isotypes and subclasses; blue – Fc-receptor binding levels; light blue – antibody functions. An asterisk indicates a global p-values obtained by non-parametric combination (Friedman test p-values for partial tests within each feature type, and Fisher's method for combination, *$p < 0.05$).
**B,C)** Heatmap showing the Spearman correlation coefficients between increases in antibody levels between day -1 and day 3 (y-axis) and corresponding donor CCP Spike-specific antibody features (x-axis) in patients with high (B) and low (C) levels of pre-existing Spike IgG1 antibody.
**D)** Heatmap depicting the difference of the Spearman correlation (donor CCP vs. day 3 – day-1) between the S_IgG1 high group and S_IgG1 low group.
**E)** Bar plot representing the statistical significance by permutation testing of the median difference between CCP features and patient trajectories in High and Low pre-existing IgG1 Spike patients. The Color represents the absolute value of the median difference and the height of the bar represents the p-value determined by the permutations test.
**F)** Bar plots of Non-parametric Combination (NPC) test on each features in donor CCP in High pre-existing S IgG1 (top) and Low pre-existing S IgG1 (bottom) individuals. Color represents the strength of the median Spearman correlation of each CCP feature in the respective High and Low pre-existing S IgG1 groups.
